## Supplementary figures and images for "Gasdermin D-Mediated Neutrophil Pyroptosis drives Inflammation in Psoriasis"

### Supplementary Figures 1-2

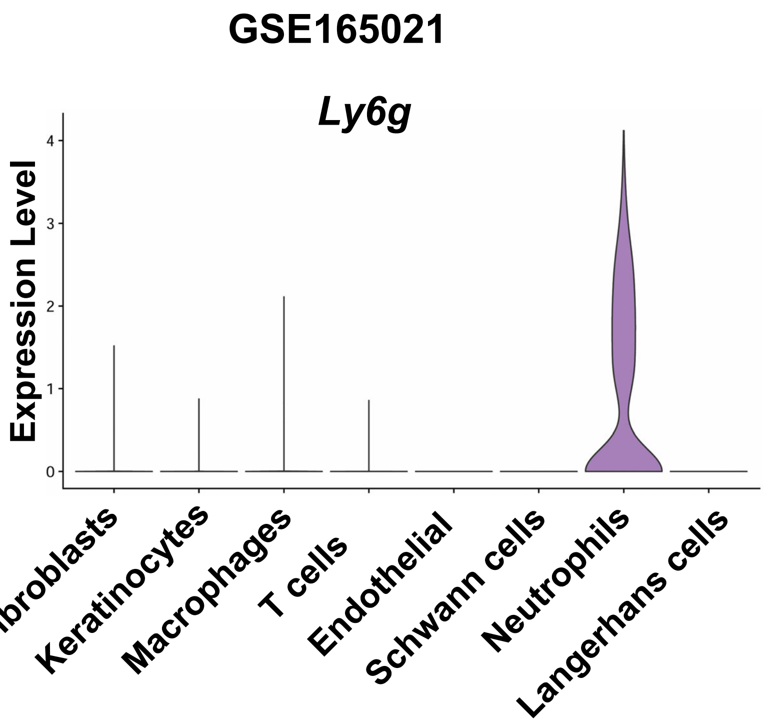

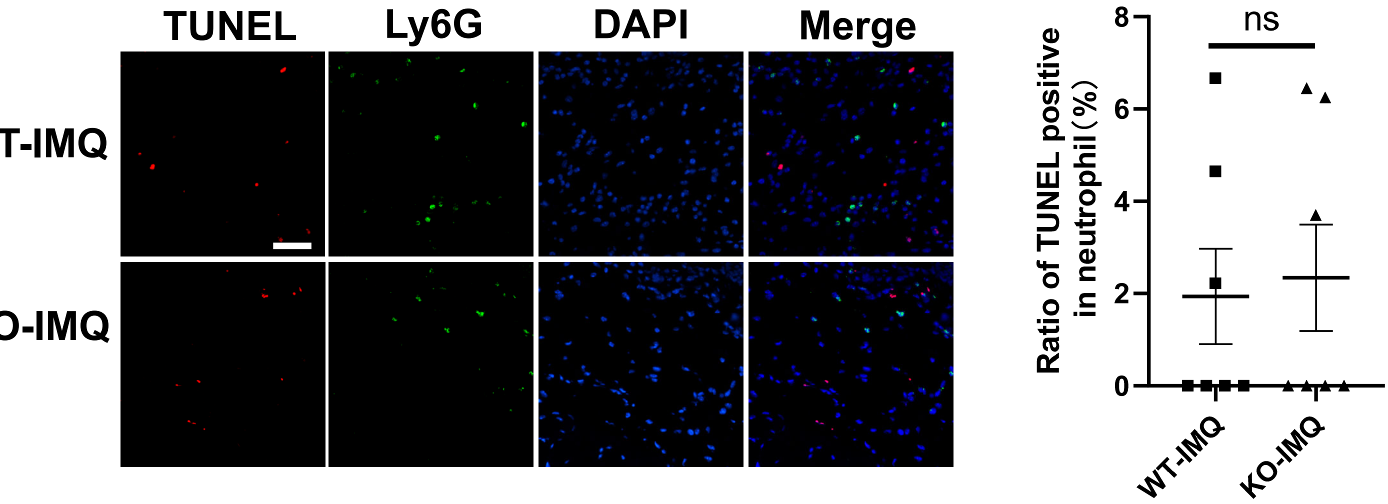

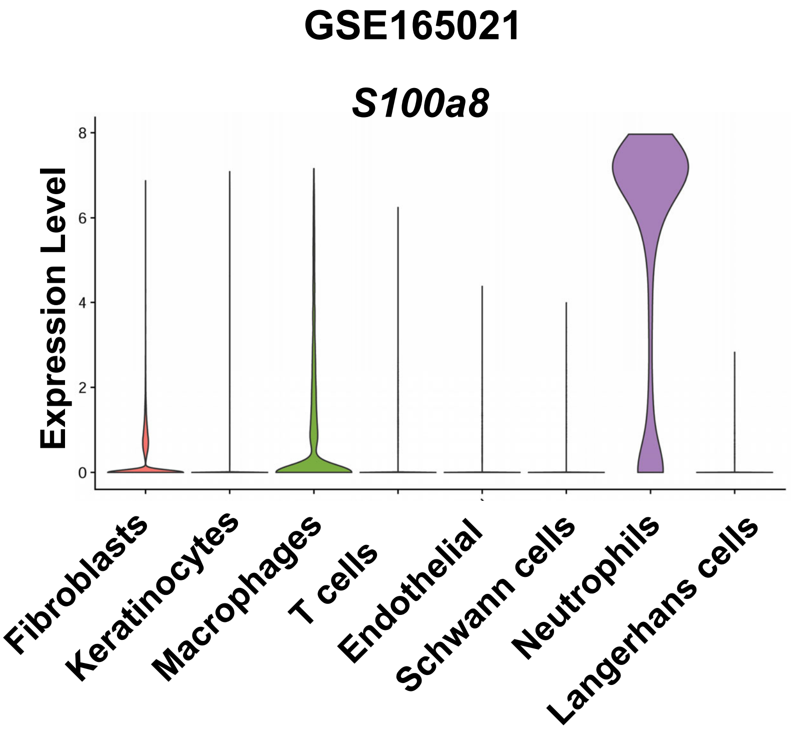

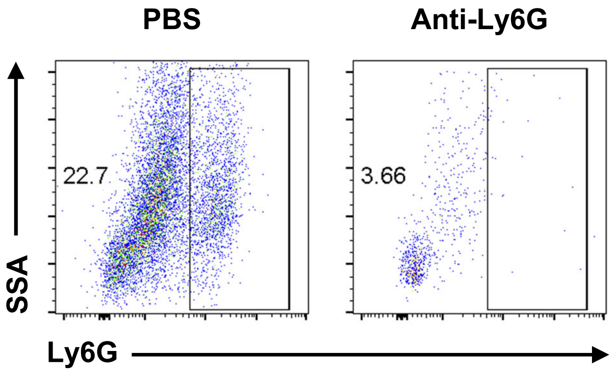

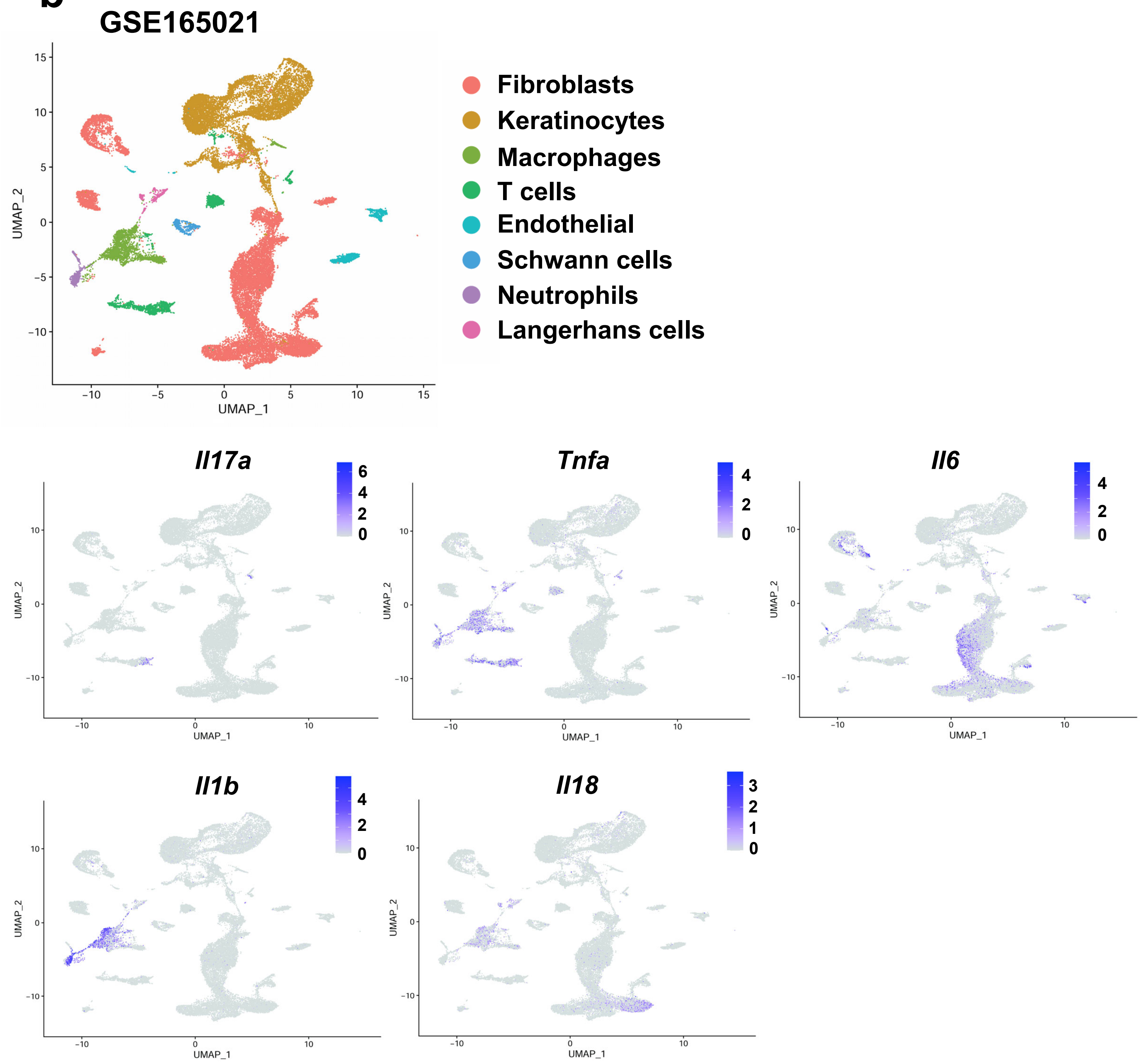
